## Supplemental Documents for "Predicting patients with false negative SARS-CoV-2 testing at hospital admission: A retrospective multi-center study"

| Supplementary Table 1. Demographic and clinical patient characteristics in cohort by COVID testing status | | | | | | |
| --- | --- | --- | --- | --- | --- | --- |
|  | **Training Dataset** | | **Testing Dataset** | |  | |
|  | **COVID +/0** | **COVID -/0** | **COVID -/+** | **COVID -/-** |  | |
|  | **N = 2,666^1^** | **N = 25,282^1^** | **N = 61^1^** | **N = 3,450^1^** | p-value^1^ | |
| **Demographics** | | | | | | |
| Age, median (IQR), years | 66.01  (52.03, 79.99) | 59.85  (37.88, 75.05) | 65.85  (53.85, 77.34) | 68.96  (55.75, 80.44) | | **<0.001** |
| Female, No/Total No (%) | 1,332 / 2,666  (50%) | 14,518 / 25,282 (57%) | 26 / 61  (43%) | 1,624 / 3,450 (47%) | | **<0.001** |
| Race, No/Total No (%) |  |  |  |  | | **<0.001** |
| Non-White | 1,407 / 2,666  (53%) | 8,338 / 25,282  (33%) | 29 / 61  (48%) | 954 / 3,450  (28%) | |  |
| White | 1,259 / 2,666  (47%) | 16,944 / 25,282 (67%) | 32 / 61  (52%) | 2,496 / 3,450 (72%) | |  |
| Latino, No/Total No (%) | 691 / 2,666  (26%) | 3,621 / 25,282  (14%) | 14 / 61  (23%) | 388 / 3,450  (11%) | | **<0.001** |
| Hospital, No/Total No (%) |  |  |  |  | | **<0.001** |
| Yale New Haven Hospital | 871 / 2,666  (33%) | 10,210 / 25,281 (40%) | 22 / 61  (36%) | 1,443 / 3,450 (42%) | |  |
| St Raphael’s Campus | 542 / 2,666  (20%) | 3,771 / 25,281  (15%) | 13 / 61  (21%) | 784 / 3,450  (23%) | |  |
| Bridgeport Hospital | 739 / 2,666  (28%) | 4,905 / 25,281  (19%) | 19 / 61  (31%) | 608 / 3,450  (18%) | |  |
| Greenwich Hospital | 421 / 2,666  (16%) | 2,432 / 25,281 (9.6%) | 6 / 61  (9.8%) | 189 / 3,450  (5.5%) | |  |
| Lawrence and Memorial Hospital | 76 / 2,666  (2.9%) | 3,099 / 25,281  (12%) | 0 / 61  (0%) | 341 / 3,450  (9.9%) | |  |
| Westerly Hospital | 17 / 2,666  (0.6%) | 864 / 25,281  (3.4%) | 1 / 61  (1.6%) | 85 / 3,450  (2.5%) | |  |
| **Baseline Characteristics, median (IQR)** | | | | | | |
| Systolic, mmHg | 131.00  (117.00, 148.00) | 133.00  (118.00, 152.00) | 125.00  (108.00, 147.00) | 134.00  (118.00, 154.00) | | **<0.001** |
| Diastolic, mmHg | 76.00  (65.00, 85.00) | 78.00  (68.00, 88.00) | 75.00  (66.00, 82.00) | 76.00  (66.00, 87.00) | | **<0.001** |
| Pulse, beats per minute | 93.00  (80.00, 108.00) | 88.00  (76.00, 102.00) | 90.00  (82.00, 112.00) | 89.00  (76.00, 105.00) | | **<0.001** |
| Respiratory rate, breaths per minute | 20.00  (18.00, 22.00) | 18.00  (17.00, 20.00) | 20.00  (18.00, 22.00) | 18.00  (18.00, 20.00) | | **<0.001** |
| spO2, % | 96.00  (93.00, 98.00) | 98.00  (96.00, 99.00) | 96.00  (94.00, 98.00) | 97.00  (95.00, 99.00) | | **<0.001** |
| Temperature, Fahrenheit | 98.74  (97.89, 100.38) | 98.03  (97.51, 98.60) | 98.74  (97.84, 100.26) | 98.08  (97.51, 98.74) | | **<0.001** |
| BMI, kg/m^2^ | 28.37  (24.15, 33.82) | 27.98  (23.88, 33.00) | 27.39  (24.06, 30.57) | 27.11  (23.10, 32.42) | | **<0.001** |
| **Comorbidities** | | | | | | |
| Elixhauser score, median (IQR) | 5.00  (2.00, 9.00) | 5.00  (2.00, 10.00) | 6.00  (3.00, 11.00) | 7.00  (4.00, 11.00) | | **<0.001** |
| CHF, No/Total No(%) | 644 / 2,666  (24%) | 6,261 / 25,282  (25%) | 17 / 61  (28%) | 1,179 / 3,450 (34%) | | **<0.001** |
| CPD, No/Total No(%) | 882 / 2,666  (33%) | 9,122 / 25,282  (36%) | 21 / 61  (34%) | 1,406 / 3,450 (41%) | | **<0.001** |
| Diabetes, No/Total No(%) | 1,069 / 2,666  (40%) | 17,607 / 25,282 (70%) | 35 / 61  (57%) | 1,337 / 3,450 (61%) | | **<0.001** |
| Obesity, No/Total No(%) | 853 / 2,666  (32%) | 7,471 / 25,282  (30%) | 23 / 61  (38%) | 1,084 / 3,450 (31%) | | **0.006** |
| Arrhythmia, No/Total No(%) | 1,006 / 2,666  (38%) | 9,874 / 25,282  (39%) | 26 / 61  (43%) | 1,675 / 3,450 (49%) | | **<0.001** |
| HTN, No/Total No(%) | 1,744 / 2,666  (65%) | 14,890 / 25,282 (59%) | 42 / 61  (69%) | 2,539 / 3,450 (74%) | | **<0.001** |
| Malignancy, No/Total No(%) | 305 / 2,666  (11%) | 4,014 / 25,282  (16%) | 7 / 61  (11%) | 680 / 3,450  (20%) | | **<0.001** |
| Metastasis, No/Total No(%) | 77 / 2,666  (2.9%) | 1,687 / 25,282 (6.7%) | 3 / 61  (4.9%) | 279 / 3,450  (8.1%) | |  |
| Alcohol abuse, No/Total No(%) | 271 / 2,666  (10%) | 4,277 / 25,282  (17%) | 7 / 61  (11%) | 525 / 3,450  (15%) | | **<0.001** |
| Drug abuse, No/Total No(%) | 258 / 2,666  (9.7%) | 4,929 / 25,282  (19%) | 6 / 61  (9.8%) | 591 / 3,450  (17%) | | **<0.001** |
| Stroke, No/Total No(%) | 155 / 2,666  (5.8%) | 1,158 / 25,282 (4.6%) | 7 / 61  (11%) | 220 / 3,450  (6.4%) | |  |
| TIA, No/Total No(%) | 55 / 2,666  (2.1%) | 511 / 25,282  (2.0%) | 2 / 61  (3.3%) | 107 / 3,450  (3.1%) | | **<0.001** |
| HIV, No/Total No(%) | 40 / 2,666  (1.5%) | 340 / 25,282  (1.3%) | 1 / 61  (1.6%) | 72 / 3,450  (2.1%) | | **0.009** |
| **Laboratory Results** | | | | | | |
| Sodium, mmol/L | 137.00  (134.00, 140.00) | 138.00  (135.00, 140.00) | 138.00  (136.00, 143.00) | 138.00  (135.00, 140.00) | | **<0.001** |
| Potassium, mmol/L | 4.00 (3.70, 4.40) | 4.00 (3.70, 4.40) | 4.10 (3.70, 4.30) | 4.10 (3.70, 4.50) | | **<0.001** |
| Bicarbonate, mmol/L | 24.40  (22.00, 27.00) | 24.00  (22.00, 27.00) | 24.00  (22.00, 26.25) | 24.55  (22.00, 27.20) | | **<0.001** |
| BUN, mg/dL | 18.00  (12.00, 30.00) | 17.00  (12.00, 27.00) | 20.00  (13.00, 34.00) | 20.00  (14.00, 31.00) | | **<0.001** |
| Creatinine, mg/dL | 1.00  (0.78, 1.47) | 0.97  (0.74, 1.37) | 1.08  (0.81, 1.60) | 1.03  (0.80, 1.47) | | **<0.001** |
| Chloride, mmol/L | 100.00  (97.00, 104.00) | 102.00  (98.00, 105.00) | 102.00  (98.00, 107.00) | 101.00 (98.00, 105.00) | | **<0.001** |
| Glucose, mmol/L | 122.00  (104.00, 162.00) | 117.00  (99.00, 149.00) | 125.00  (109.00, 161.00) | 124.00  (105.00, 160.00) | | **<0.001** |
| Hemoglobin, g/dL | 12.90  (11.50, 14.30) | 12.50  (10.90, 13.90) | 12.50  (10.50, 14.00) | 12.30  (10.40, 13.90) | | **<0.001** |
| Platelet count, x10^9^/L | 204.00  (159.00, 263.00) | 231.00  (180.00, 291.00) | 234.00  (190.00, 327.00) | 236.00  (180.00, 301.25) | | **<0.001** |
| WBCC, x10^3^ mm^3^ | 6.80  (5.10, 9.40) | 9.20  (6.90, 12.10) | 8.60  (5.80, 11.70) | 9.50  (7.00, 12.90) | | **<0.001** |
| % Lymphocyte, % | 15.10  (9.40, 22.10) | 16.50  (10.00, 24.20) | 13.90  (10.20, 17.80) | 13.50  (7.70, 21.50) | | **<0.001** |
| Anion Gap. mmol/L | 14.00  (12.00, 16.00) | 13.00  (10.00, 15.00) | 14.00  (12.00, 16.00) | 13.00  (10.00, 16.00) | | **<0.001** |

Supplementary Figure 1A. Closed form equation and coefficients for regression coefficients model to predict probability of having a COVID positive test

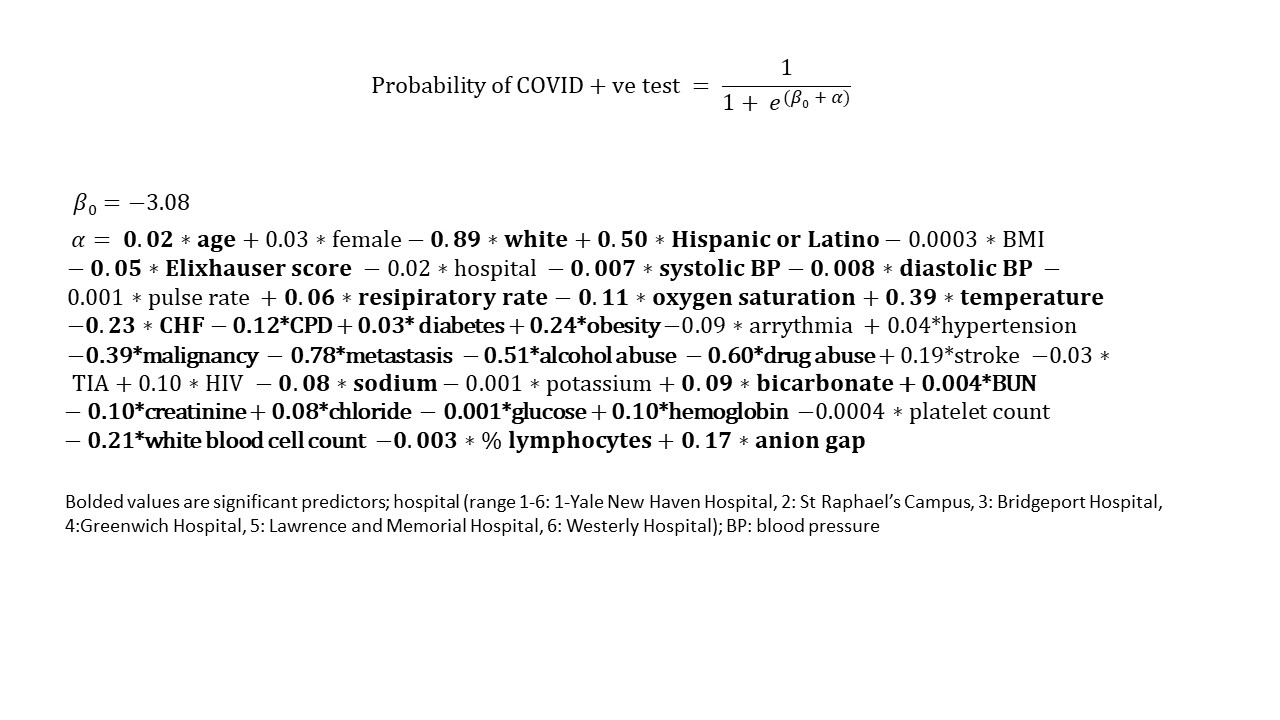

Supplementary Figure 1B. Z-scores of model covariates

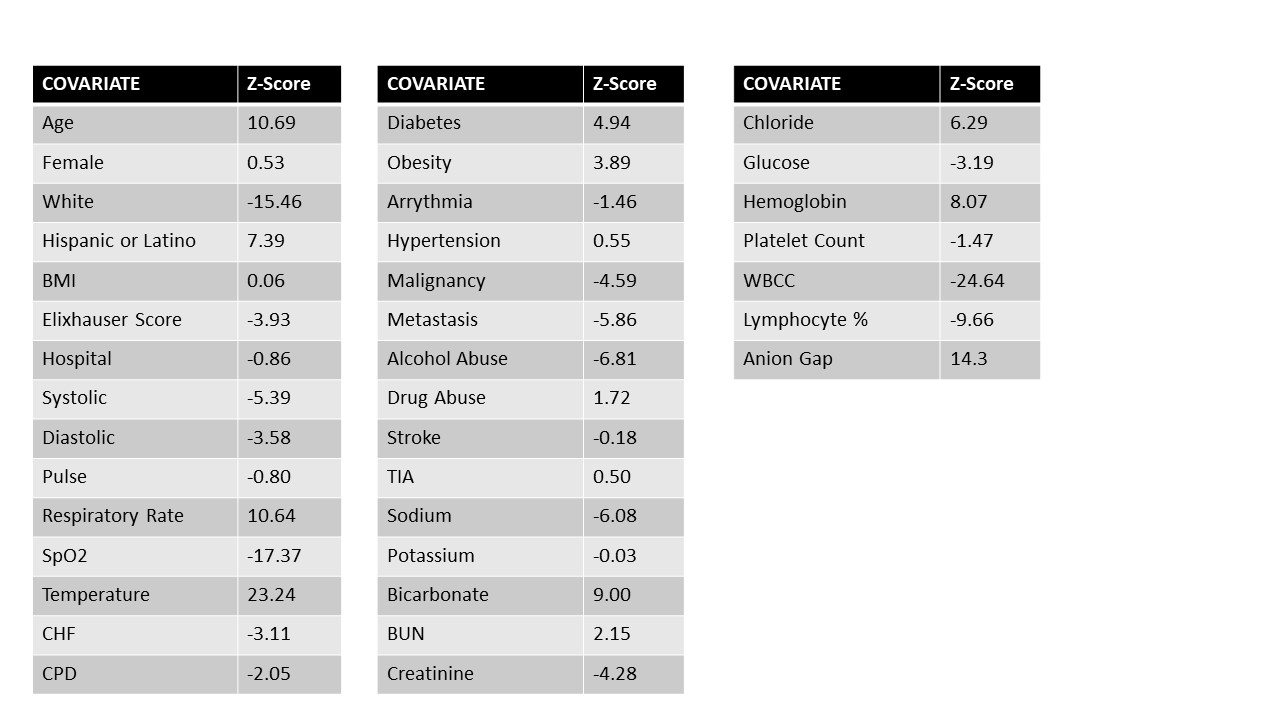

Supplementary Figure 2. Precision/Recall Curve to detect COVID positivity among those who had a negative COVID test on admission and were retested

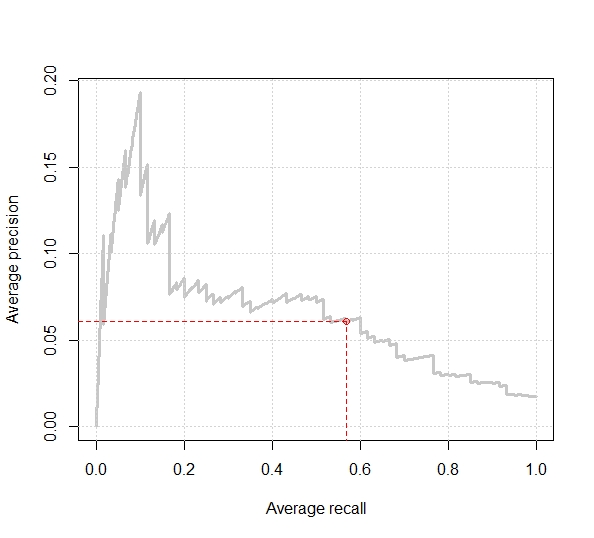

Precision is the positive predictive value of having a COVID positive test and recall is the sensitivity of having a positive COVID test. Sensitivity in our testing cohort is 0.57 and the positive predictive value is 0.61 as can be seen on this graph.
